# Impact of city and residential unit lockdowns on prevention and control of COVID-19

**DOI:** 10.1101/2020.03.13.20035253

**Authors:** Peng Shao

## Abstract

With respect to the asymptomatic transmission characteristics of the novel coronavirus that appeared in 2019 (COVID-19), a susceptible-asymptomatic-infected-recovered-death (SAIRD) model that considered human mobility was constructed in this study. The dissemination of COVID-19 was simulated using computational experiments to identify the mechanisms underlying the impact of city and residential lockdowns on controlling the spread of the epidemic. Results: The implementation of measures to lock down cities led to higher mortality rates in these cities, due to reduced mobility. Moreover, implementing city lockdown along with addition of hospital beds led to improved cure and reduced mortality rates. Stringent implementation and early lockdown of residential units effectively controlled the spread of the epidemic, and reduced the number of hospital bed requirements. Collectively, measures to lock down cities and residential units should be taken to prevent the spread of COVID-19. In addition, medical resources should be increased in cities under lockdown. Implementation of these measures would reduce the spread of the virus to other cities and allow appropriate treatment of patients in cities under lockdown.

## 1. Introduction

A novel coronavirus was identified as the cause of viral pneumonia outbreaks in Wuhan, China, in 2019. The World Health Organization officially termed this novel virus COVID-19 and publicly announced this on February 11, 2020. Patients infected with COVID-19 can be asymptomatic carriers[1] and infect others despite having no obvious symptoms. Because many infected patients are not or mildly symptomatic, identification of the chain of transmission and contact tracing have become extremely complicated [2]. Some studies of COVID-19 have used mathematical models and computer technologies to simulate and predict COVID-19 transmission[3-5]. For example, public health investigators evaluated the COVID-19 epidemic in China during the initial phase[6].

The Chinese central government implemented lockdown measures in Wuhan and other cities in Hubei Province to quarantine core areas of the COVID-19 outbreak on January 23, 2020. Two weeks later, measures to lock down residential units started to be implemented in most cities in China. However, the virus has spread across national borders to several other countries[7]. After the Chinese government decided to lock down cities in January, questions arose regarding the reasons behind a nationwide lockdown of residential units that was implemented in February, the impact of these measures on the control of COVID-19 dissemination, and whether they could provide any significant information that might help other countries faced with a similar situation. Here, computational experiments were conducted to identify the mechanisms underlying the impact of the lockdown of cities and residential units on the prevention and control of the epidemic.

## 2. Background

The lockdown of core cities and the nationwide lockdown of residential units were important measures adopted by the Chinese government to manage the spread of the epidemic. Lockdown measures for COVID-19 epidemic regions were first implemented in Wuhan City, Hubei Province, on January 23, 2020, followed by the progressive lockdown of other cities in the Hubei Province. By February 7, 2020, the number of new confirmed and suspected cases gradually began to decline in China (Figure 1A and C). From February 8, 2020, most cities across China had also implemented measures to lockdown residential units, and only one person from each household was permitted to purchase essential supplies every two or three days. Residents were required to show an “enter and exit pass” when entering or leaving their residence, and body temperature had to be checked at both entry and exit.A sharp increase in the number of confirmed infections on February 13 (Figure 1A and D) was due to adopting the recommendations in the 5^th^ edition of the Diagnostic and Treatment Plan issued by the National Health Commission of China, which stated that a clinical diagnosis could serve as a basis for confirming a diagnosis.

**Figure 1.**
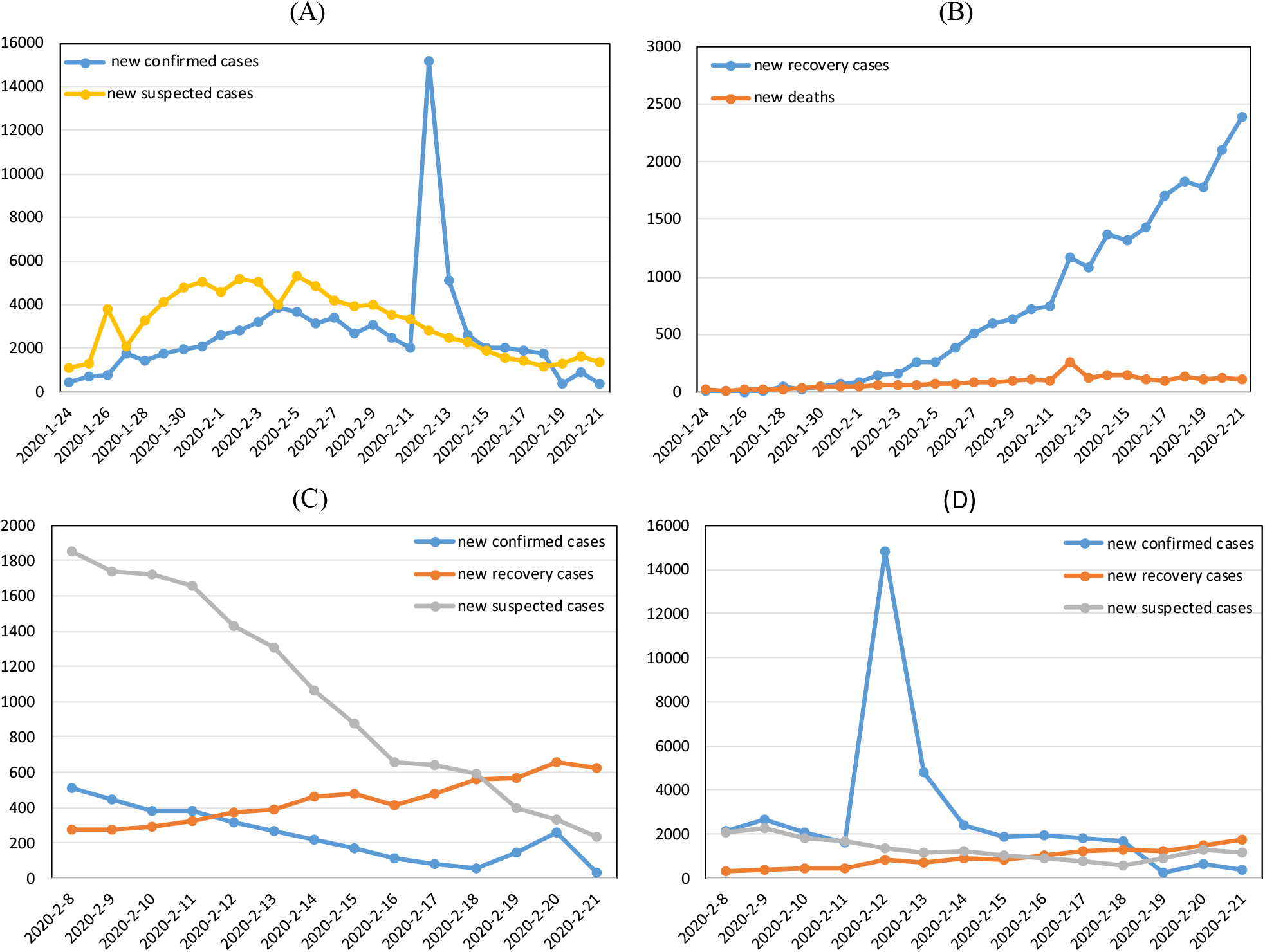
Trends in COVID-19 development between February 8, 2020 and February 21, 2020 in mainland China. (A) Numbers of new confirmed and suspected cases and (B) new recovered cases and deaths per day in mainland China. Numbers of new cases per day outside (C) and within (D) Hubei Province.

Medical resources are important factors that can determine the capacity of the core epidemic regions to provide treatments to patients. The 2019 China Statistical Yearbook states that the number of hospital beds per thousand people ranges from 4.56 to 7.19 in mainland China, and is 6.65 in Hubei Province (Table 1). However, Hubei was hit hard by the epidemic after the outbreak. A large number of confirmed infections and a limited number of hospital beds led to the failure of timely treatment for many patients, which resulted in severe illness and death.

**Table 1.**
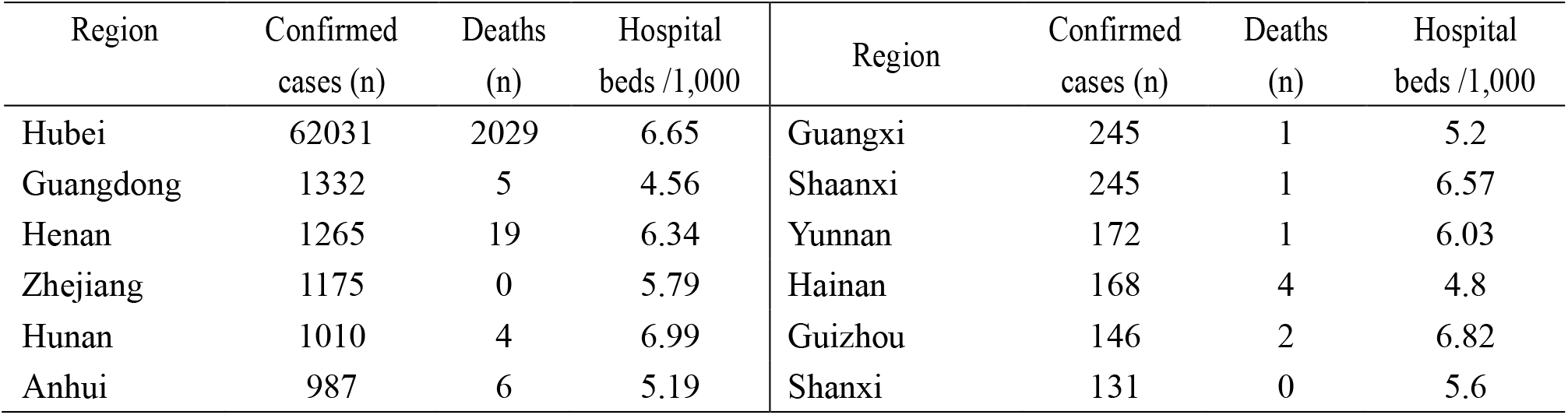
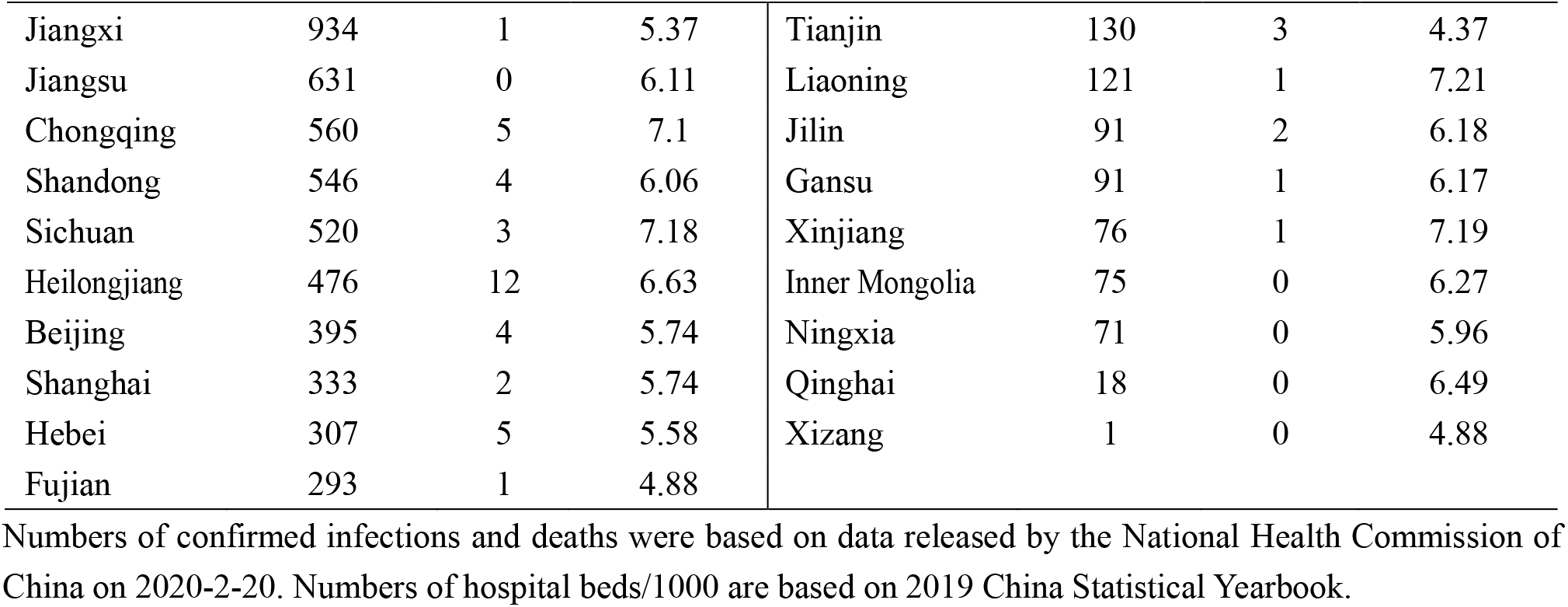
Epidemic situation and medical resources.

| Region | Confirmed cases (n) | Deaths (n) | Hospital beds /1,000 | Region | Confirmed cases (n) | Deaths (n) | Hospital beds /1,000 |
| --- | --- | --- | --- | --- | --- | --- | --- |
| Hubei | 62031 | 2029 | 6.65 | Guangxi | 245 | 1 | 5.2 |
| Guangdong | 1332 | 5 | 4.56 | Shaanxi | 245 | 1 | 6.57 |
| Henan | 1265 | 19 | 6.34 | Yunnan | 172 | 1 | 6.03 |
| Zhejiang | 1175 | 0 | 5.79 | Hainan | 168 | 4 | 4.8 |
| Hunan | 1010 | 4 | 6.99 | Guizhou | 146 | 2 | 6.82 |
| Anhui | 987 | 6 | 5.19 | Shanxi | 131 | 0 | 5.6 |
| Jiangxi | 934 | 1 | 5.37 | Tianjin | 130 | 3 | 4.37 |
| Jiangsu | 631 | 0 | 6.11 | Liaoning | 121 | 1 | 7.21 |
| Chongqing | 560 | 5 | 7.1 | Jilin | 91 | 2 | 6.18 |
| Shandong | 546 | 4 | 6.06 | Gansu | 91 | 1 | 6.17 |
| Sichuan | 520 | 3 | 7.18 | Xinjiang | 76 | 1 | 7.19 |
| Heilongjiang | 476 | 12 | 6.63 | Inner Mongolia | 75 | 0 | 6.27 |
| Beijing | 395 | 4 | 5.74 | Ningxia | 71 | 0 | 5.96 |
| Shanghai | 333 | 2 | 5.74 | Qinghai | 18 | 0 | 6.49 |
| Hebei | 307 | 5 | 5.58 | Xizang | 1 | 0 | 4.88 |
| Fujian | 293 | 1 | 4.88 |  |  |  |  |

Numbers of confirmed infections and deaths were based on data released by the National Health Commission of China on 2020-2-20. Numbers of hospital beds/1000 are based on 2019 China Statistical Yearbook.

## 3. Model

Based on the SEIR model and considering asymptomatic COVID-19 carriers, deaths and the mobility of individuals, we constructed a model that included S, A, I, R, and D states (SAIRD). The status of individuals in the SAIRD model primarily included the following transition modes:

An S individual who contacts an I individual or an A carrier will have the probability of becoming an A carrier.

An A carrier might transition to the I state after the incubation period.

An I individual will have a specific likelihood of being quarantined and treated, thus transitioning into the R state.

An I individual who has not been effectively treated after a specific period might transition into the D state.

In addition, the movements of individuals in I and A states across geographic areas during the COVID-19 epidemic resulted in rapid spread, resulting in epidemics in other cities. Therefore, the spread of the epidemic caused by human movement was simulated using computational means in the present study.

## 4. Experimental design

The simulation experiments were conducted using MATLAB R2017a and included simulations of locked down cities (LC) and locked down residential units (LU). Table 2 shows the parameter settings.

**Table 2.**
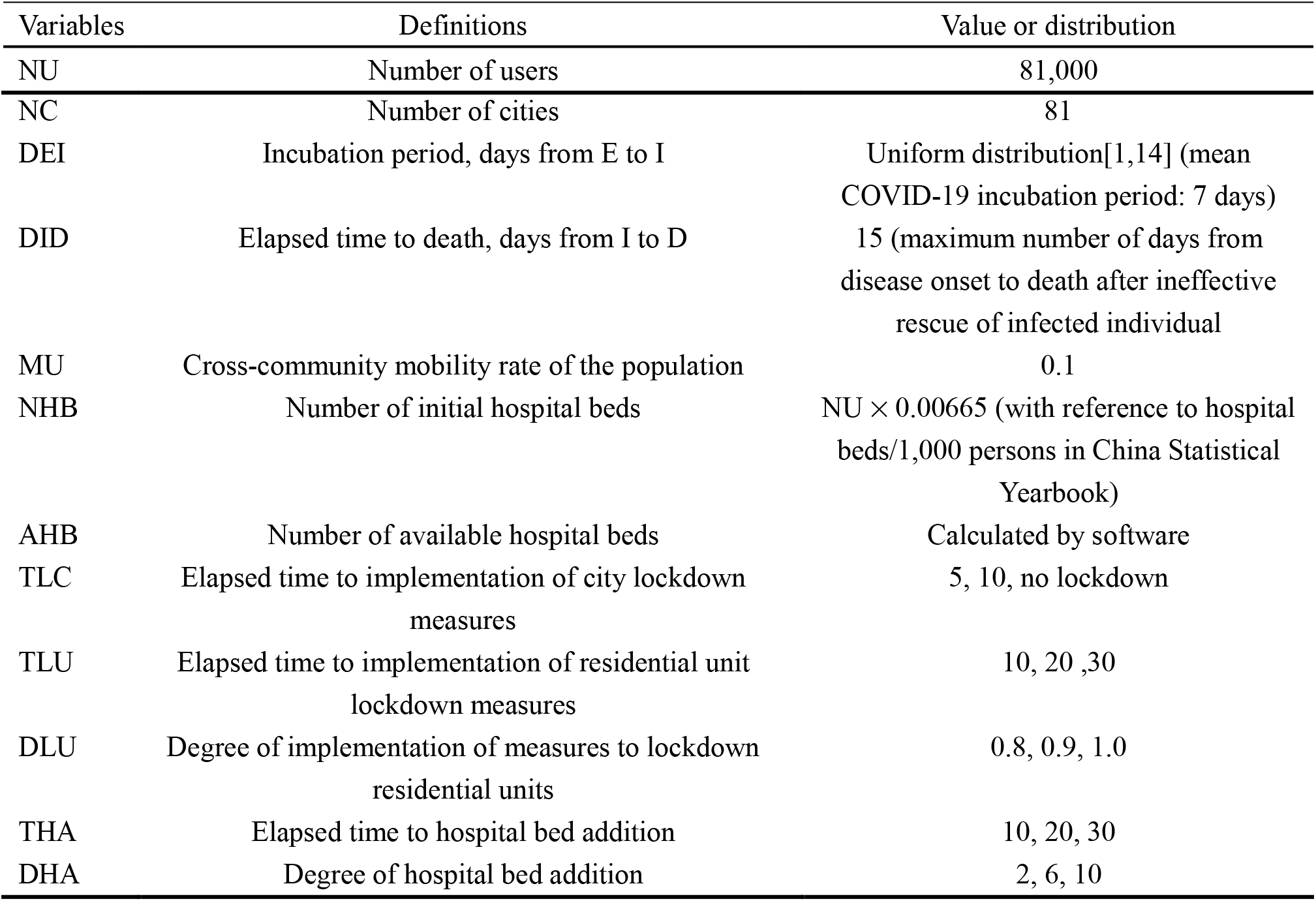
Parameter settings.

| Variables | Definitions | Value or distribution |
| --- | --- | --- |
| NU | Number of users | 81,000 |
| NC | Number of cities | 81 |
| DEI | Incubation period, days from E to I | Uniform distribution[1,14] (mean COVID-19 incubation period: 7 days) |
| DID | Elapsed time to death, days from I to D | 15 (maximum number of days from disease onset to death after ineffective rescue of infected individual) |
| MU | Cross-community mobility rate of the population | 0.1 |
| NHB | Number of initial hospital beds | $NU \times 0.00665$ (with reference to hospital beds/1,000 persons in China Statistical Yearbook) |
| AHB | Number of available hospital beds | Calculated by software |
| TLC | Elapsed time to implementation of city lockdown measures | 5, 10, no lockdown |
| TLU | Elapsed time to implementation of residential unit lockdown measures | 10, 20, 30 |
| DLU | Degree of implementation of measures to lockdown residential units | 0.8, 0.9, 1.0 |
| THA | Elapsed time to hospital bed addition | 10, 20, 30 |
| DHA | Degree of hospital bed addition | 2, 6, 10 |

Computational experiments were conducted in two-dimensional space with a population size of NU. In the initial phase, NU was randomly and uniformly distributed in NC; that is, the number of individuals in each city was NU/NC. One individual in a major city was randomly selected as the first infected person. The simulation program ran 200 timeframes, during which individuals might undergo spatial movement and a state change at any time frame (for example, daily). First, without considering the status of city and residential unit lockdowns, individuals moved to neighboring cities at a probability of the across-community mobility rate of the population (MU). Secondly, individuals in all cities either changed or maintained their original states based on the rules of the SAIRD model.

## 5. Simulation

### 5.1 Simulation of a city under lockdown

Figure 2 shows differences in the ratios of individuals in the five states in the core locked down cities and the ratios of infected users in each city. With respect to timing, if a city was locked down after an infected user was confirmed and given no residential unit lockdown and no increase in medical resources, the ratio of individuals in S state would be 0% at steady state; that is, all individuals would be infected. This was without regard to whether the city was under early (Figure 2C), late (Figure 2B), or no lockdown (Figure 2A). Regarding variance in the ratios of D individuals among all cities, the earlier a city was locked down (Figure 2F, TLC = 5), the greater the variance in the proportions of D individuals state across all 81 cities (VARD ∼ 0.05). This indicated that the number of deaths in the core cities that were under lockdown was significantly higher than in other cities. By comparison, variance in the proportions of D individuals across the 81 cities was lower when no cities were locked down (VARD of ∼ 0.02; Figure 2D). Overall, the implementation of measures to place cities under lockdown could not reduce the ratios of infected individuals in the SAIRD model. The implementation of measures to lock down cities would result in larger variances in the number of deaths between cities with and without lockdown. These findings were consistent with the perspective of Li et al. (2020), who reported that the stringent lockdown policy applied to Hubei Province had suspended a nationwide outbreak of COVID-19[8]. However, mortality rates in lockdown cities were higher because infected individuals in core cities were unable to move to other cities.

**Figure 2.**
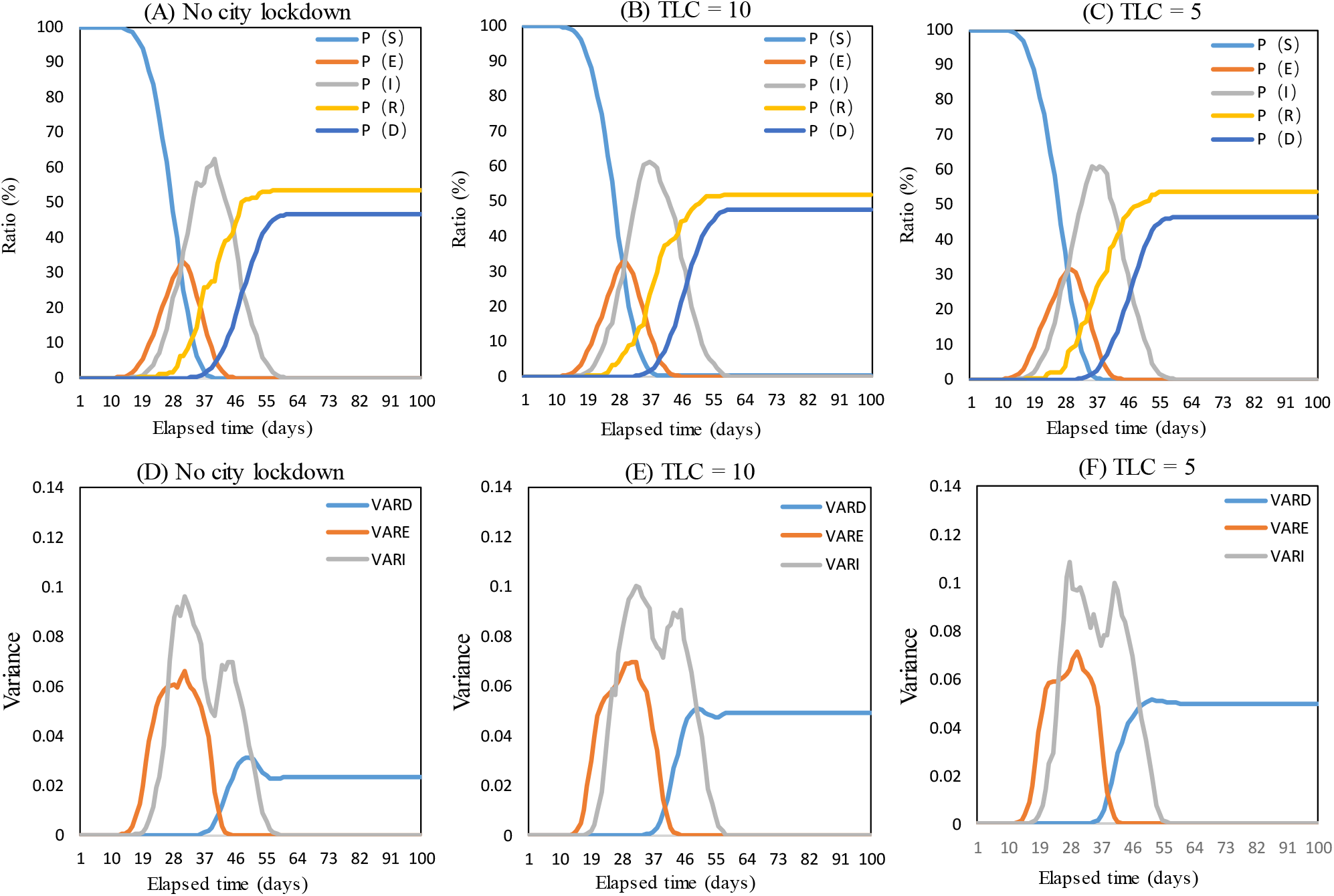
Impact of elapsed time to city lockdown on COVID-19 spread.

We investigated the impact of adding hospital beds on the spread of the epidemic as follows. TLC and THA were both set at 10. We then examined the proportions of individuals in the five states when the hospital beds were increased 2-(DHA=2), 6-(DHA= 6), and 10-(DHA = 10) fold (Figure 3). The addition of hospital beds increased cure rates and reduced the mortality rates, but could not significantly reduce infection rates. When fewer hospital beds were added (Figure 3A, DHA = 2), the ratios of individuals in the R and D state were about 55% and 45%, respectively, at steady state. When more hospital beds were added (Figure 3C; DHA = 10), the ratios of individuals in the R and D states were about 85% and 15%, respectively, at steady state.

**Figure 3.**
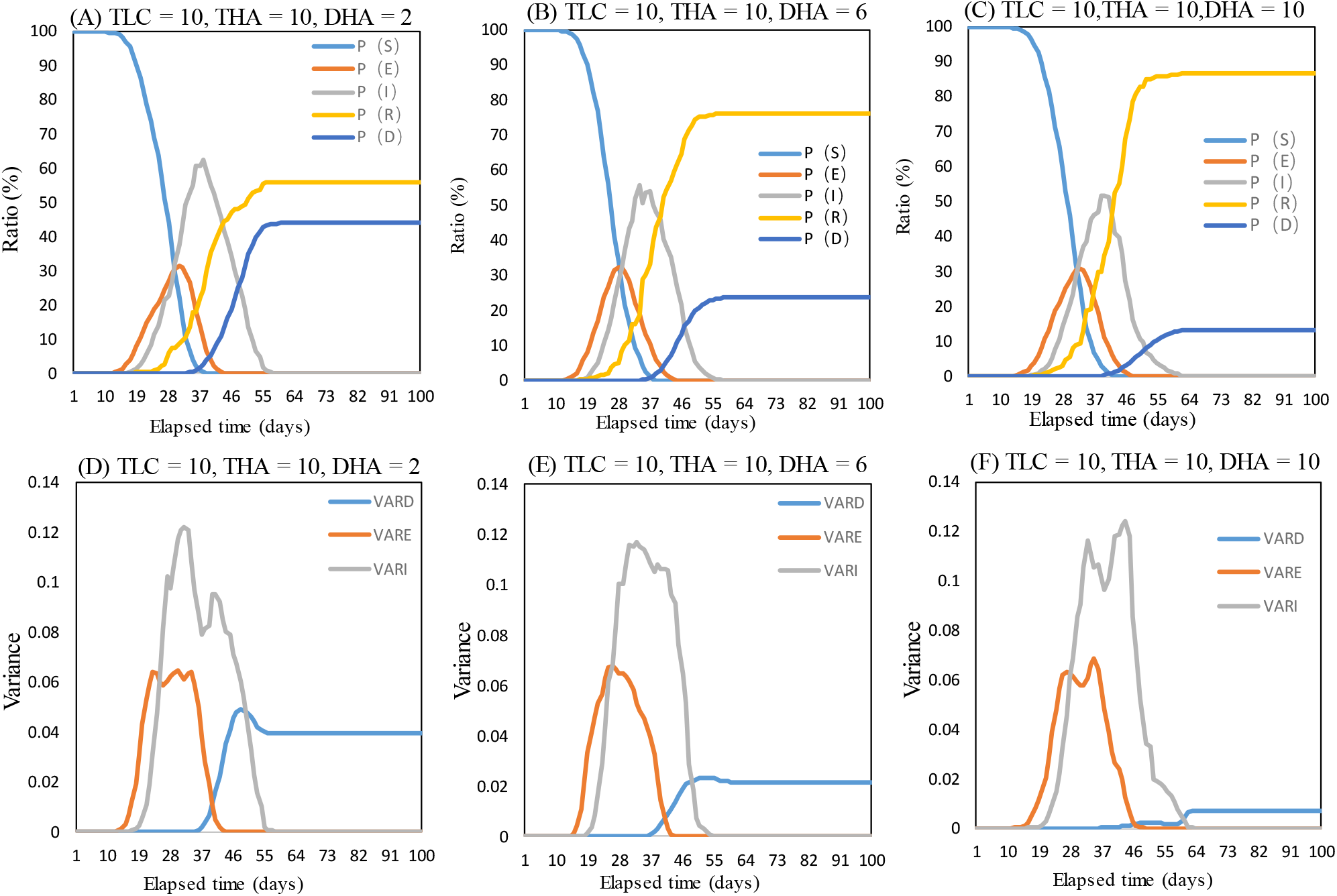
Impact of city lockdown and hospital bed addition on COVID-19 spread.

We examined variances in the numbers of individuals in D state across 81 cities. Variance in the proportions of deaths across these cities at a steady state was larger when fewer hospital beds were added (Figure 3D; DHA = 2; variance, ∼ 0.04). In contrary, variance in the proportions of deaths across these cities at a steady state was smaller when more hospital beds were added to core cities (Figure 3F; DHA = 10; variance, ∼ 0.005). Overall, after implementing measures to lock down the cities, adding more hospital beds effectively improved cure rates and reduced mortality rates. Moreover, adding more hospital beds to core cities resulted in a low variance in the ratios (%) of deaths in the 81 cities. This indicated that increasing the amount of medical resources (number of hospital beds) could effectively reduce mortality rates in cities under lockdown.

### 5.2 Simulation of residential unit lockdown

Figure 4 shows the proportions of individuals in the five states in the SAIRD model and the number of available hospital beds after implementing measures to lock down residential units. The DLU was set at 0.9 and the TLU at 10, 20, and 30. When residential units were placed under lockdown sooner (Figure 4A; TLU = 10), the proportions of individuals in S state when steady state was reached was 0.95. This indicated more uninfected, and less infected individuals. Later lockdown (Figure 4C; TLU = 30), resulted in ∼ 15% of the population being in the S state when steady state was reached. This indicated less uninfected individuals and more infected individuals. Later implementation of measures to lock down residential units (Figure 4F, TLU = 30), resulted in more I individuals, which led to a worsened bed shortage (a greater negative peak of available hospital beds). Sooner implementation of residential unit lockdown measures (Figure 4D, TLU = 10), resulted in less individuals becoming infected. Thus, a bed shortage did not arise. Overall, the lockdown of residential units effectively controlled the spread of the epidemic, particularly when implemented sooner. This was reflected by lower infection rates and a sufficient number of hospital beds.

**Figure 4.**
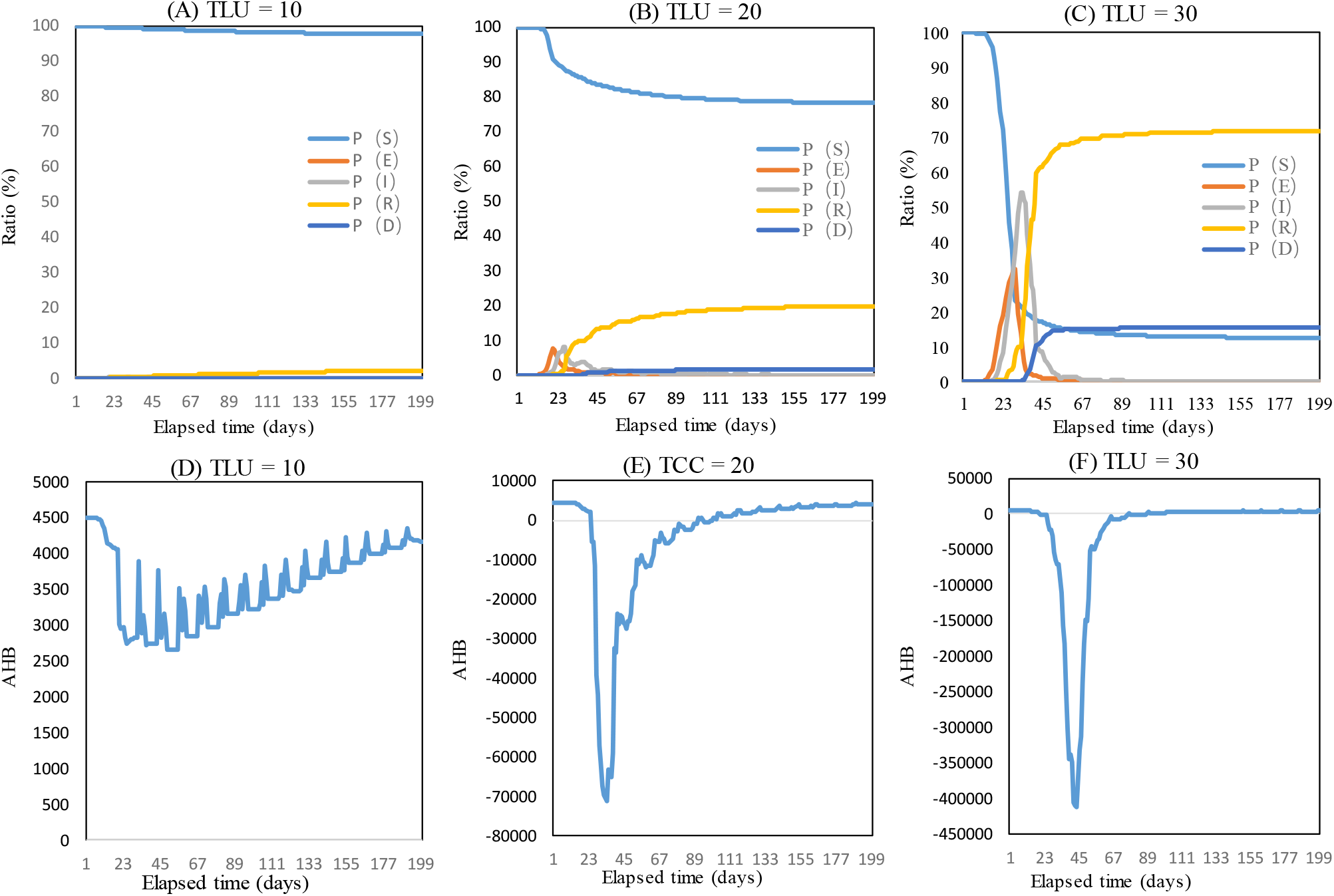
Impact of elapsed time to implementation of measures to lock down residential units on COVID-19 spread.

Further, the TLU was set at 20 and the DLU at 1.0, 0.9, and 0.8, and the proportions of individuals in the five states and the number of available hospital beds were examined (Figure 5). A higher degree of implementation of measures to lock down residential units led to lower infection rates (Figure 5A; DLU = 1.0). At steady state, the proportion of S individuals was higher, and almost 85% of the individuals were not infected. This result showed that stringent implementation of measures to lock down residential unit could effectively control the spread of the epidemic. If the DLU was relaxed (Figure 5C; DLU = 0.8), the number of infected individuals significantly increased to 50% and the hospital bed shortages were prolonged. For example, under the complete lockdown of residential units (Figure 5D; DLU = 1.0), hospital bed shortage occurred at time frames of 20 – 40. In contrast, hospital bed shortages occurred at time frames 20 – 140 under non-stringent residential unit lockdown (Figure 5F; DLU = 0.8). The area under the curve (AUC) for the number of available beds that indicated the number of hospital bed shortages in a given period was smaller for complete lockdown (Figure 5D; DLU = 1.0) compared with the non-stringent lockdown of residential units (Figure 5F; DLU = 0.8). These findings indicated that more stringent implementation of the measures to lock down residential units led to lesser hospital bed requirements.

**Figure 5.**
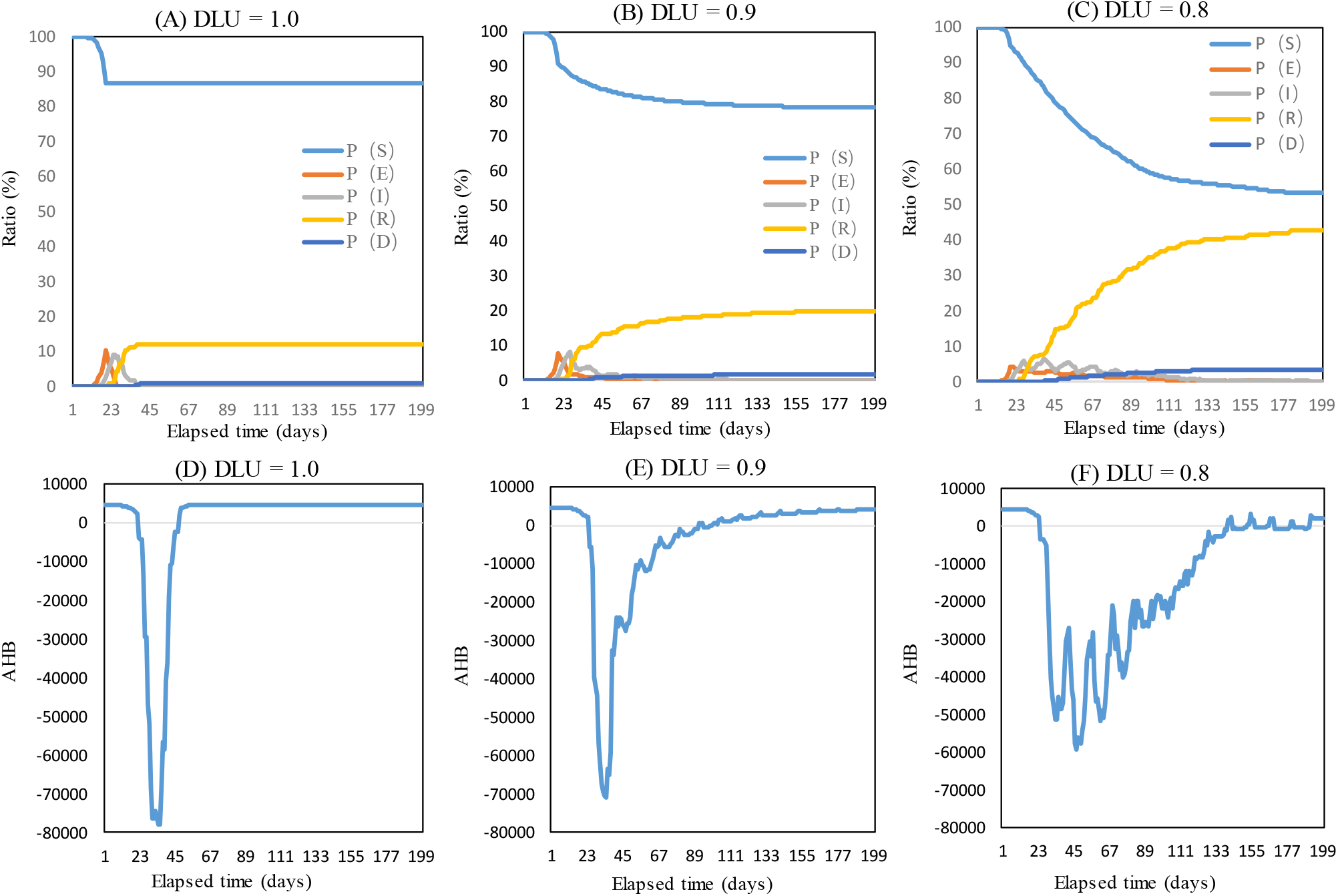
Impact of implementation of measures to lock down residential units on epidemic spread.

## 6. Conclusions

Our findings revealed that solely implementing measures to place cities under lockdown could not reduce the proportions of infected individuals in the SAIRD model and would also lead to higher mortality rates in these cities. Adding a large number of hospital beds along with city lockdown improved cure rates and reduced mortality rates. Residential unit lockdown effectively control the spread of the epidemic, and sooner implementation of this measure resulted in more effective epidemic control.

The epidemic control measures adopted by China (city and residential unit lockdown) might provide valuable information for other countries. Italy has the worst epidemic situation in Europe. From February 23, 2020, Italy imposed quarantine and lockdown measures on > 12 municipalities, banned public assembly, and closed venues such as schools and bars; these actions were similar to the situation in Wuhan, China. However, the present findings indicate that Italian government should recommend that their citizens also undergo stringent home quarantine (residential lockdown). In addition, medical resources in the affected regions should be increased as soon as possible, and specialized hospitals for patients infected with COVID-19 should be provided.

## Data Availability

Availability of all data referred to in the manuscript links below

http://www.nhc.gov.cn/xcs/yqfkdt/gzbd_index.shtml

